# Comparative evaluation of serum lithium estimation in serum samples using plain glass vial and serum clot activator vacutainer by reflectance photometry principle

**DOI:** 10.1101/2023.01.11.23284245

**Authors:** Prabesh kumar Sahu, Sibasish Sahoo, Namrata Chatterjee, Jitendra Majhi, Atanu Kumar Dutta, Kalyan Goswami, Amit Pal

## Abstract

**Background:** The collection of blood samples in different vacutainers can affect the result of serum lithium estimation due to the presence of distinct additives added in the blood collection vacutainer for enhancing the clot formation process. The presence of these additives has proven to interfere with laboratory results of biochemical analytes. In this case, both the pre-analytical and the analytical phases have a significant impact on the accuracy and precision of the test result. Thus, it has become a challenge for the lab physician to estimate lithium in any clinical laboratory setup.

**Materials and Methods:** In total, 100 patient samples were collected and paired into clot activator vacutainers and plain glass vials. Both the paired collecting tubes after centrifugation were processed immediately via Vitros 4600 analyzer working on the principle of reflectance photometry. Both the paired tubes were stored at 2-8 degrees centigrade and were further analyzed, at 24 and 48 hours respectively, from the time of their collection. The statistical analysis was done in IBM® SPSS® software version 23.

**Results:** There was a significant difference in the lithium values obtained from clot activator vacutainers in comparison to glass vial. The difference between the measurements of the lithium levels was statistically tested by ‘t’ test where it was observed that the lithium measurements by the two different collecting tubes differed significantly with a Confidence Interval of 0.03 to 0.06 implying the level of agreement of both the methods are not similar. But there was no statistical difference in the lithium values estimated at 24 hours and 48 hours of collection.

**Conclusion:** In this study, lithium levels measured by clot-activated vacutainers are found to be low as compared to lithium levels measured through glass vials. There is no apparent change in lithium values obtained at 1hr, 24hrs & 48hrs from their collection, when measured from the glass vials and the clot-activated vacutainers respectively, by the Vitros 4600 dry chemistry analyzer.

**Summary Statement:** Serum Lithium estimation in clincal laboratory may be done different methodologies, but the result may vary due to change in principle of investigation and the pre analytical and analytical interferences.

## Introduction

Prescription of oral lithium in clinical practice started with the treatment of gout [1] but with subsequent time course, it has proven to be a potent drug in the treatment of bipolar mood disorder. Due to its narrow therapeutic window, the therapeutic drug monitoring (TDM) of Lithium (Li) has to be measured at designated intervals to maintain a constant concentration in a patient’s bloodsera, for optimizing individual dosage regimens [2].To condemn the risk of severe toxicity effects of high doses of lithium like tremors, renal failure, coma or even death, lithium blood levels must be monitored continuously during the phase of treatment. To be in steady-state concentrations and to achieve the remission of symptoms, the lithium concentration in the patients undergoing lithium therapy should be monitored at an interval of every 1 to 3 months.[3]

The currently recognized standard draw time for estimation of lithium serum concentrations is at least 10 to 12 hours after the administration of the evening dose on a twice-daily dosing regimen.[4] Serum and plasma are the main specimens used in laboratories for the determination of lithium levels. Accurate monitoring of lithium blood concentrations and complete evaluation of the method used for it requires consideration of preanalytical factors such as specimens’ type and aspect, intra, and inter-individual variations. Analytical factors such as specificity, selectivity, linearity, and total imprecision must be taken into account for good clinical laboratory practices.[5]

There are reports in the literature that proclaim a positive interference by silicone which is present as an added additive by the vacutainer manufacturer to stabilize clot formation in clot activator (red capped) vacutainers where they interact with ions selective membrane (ISE) leading to increased voltage signal thus, falsely elevating lithium levels [6]. This may give on to false alarms concluding to mismanagement in the patients undergoing Li therapy and monitoring.

Optical techniques such as the ion selective electrode method, fluorimetry and spectrophotometry have been employed in blood lithium measurement by several studies. Presumably, there is no study available on the evaluation of concentration and stability on storage for Li estimation in different specimen types on the VITROS 4600 dry chemistry analyzer based, on the reflectance photometry principle. In this line of work, Li estimation in sera from vacutainers with clot activator will be compared with that of Li estimation in sera from glass vacutainers.

## Materials and Methods

During the study period, a total of 100 patients attended the outpatient department of All India Institute of Medical Sciences, Kalyani and all were included in this study. They were undergoing lithium therapy and were advised for therapeutic monitoring of serum lithium to the Biochemistry clinical laboratory, so no exclusion of patients was made. A volume of 5 ml of blood was drawn from the antecubital vein using a 5 ml syringe following all antiseptic precaution measures. Immediately after collection, 2.5 ml of blood was equally transferred into the plain glass vials and the clot activator vacutainers respectively (Table 1). After allowing both vacutainers to clot for 20-30 minutes, the vacutainers were centrifuged at 3500 rpm for 10 mins.

**Table 1:** Shows the two types of collection tube with presences of additives.

| Sl.No. | Types of Blood collection vial | Amount of blood collected in vial | Any additives presenting the collecting blood vial |
| --- | --- | --- | --- |
| 1 | clot activator vacutainer | 2.5 ml | Silicon powder for enhancing clot formation |
| 2 | Plain Glass vial | 2.5 ml | No additives |

After proper centrifugation, both the collection tubes were immediately processed by Vitros 4600 analyzer in the laboratory within 1 hour of its collection. After processing, both the collection tube were stored at 2°C–8°C and were again processed in the VITROS 4600 at 24 hours and 48 hours of sample collection. Prioritizing the quality control, procedures were followed as per the protocol of the clinical Biochemistry Lab.

For the study, an ethical clearance (IEC/2022/47) was obtained from the institute’s ethics committee. Proper informed consent was taken from the patients and confidentiality was maintained during this study. The statistical analysis was done by IBM® SPSS® software version 23.

## Results

Of the 100 blood samples collected and analyzed in pairs, it was observed that blood sample collected in clot activator vacutainer had lower lithium values in comparison to the Glass vial collection method. The mean baseline ‘lithium’ value by the glass vial collection method was 0.61 and by the Clot vacutainer collection method was 0.58

The immediate observation of lithium levels by both the collecting tubes was made within the 1st hour of blood collection. The mean of the difference of the serum Lithium levels by the two collection methods was found to be 0.04 with a standard deviation of 0.06 (Table 2). The ‘t’ test for the difference between the measurements of the lithium levels was statistically tested, where in it was observed that the lithium measurements by the two methods differed significantly with a Confidence Interval of 0.03 to 0.06 implying the level of agreement of both the methods are not similar (Table 2).

**Table 2:**
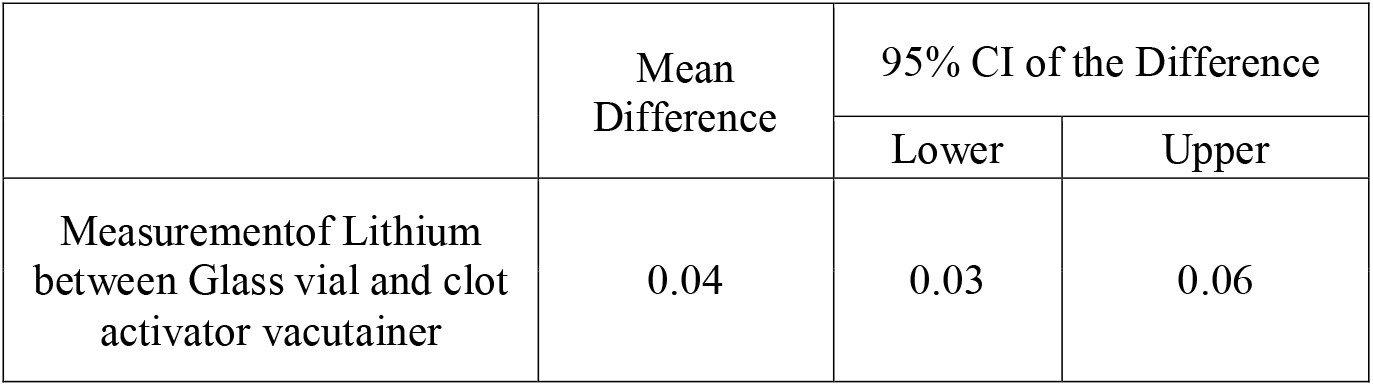
Shows the t test for the difference between the measurements of the lithium levels by glass vials and clot vials with a Confidence Interval of 0.03 to 0.06 implying the level of agreement of both the methods are not similar.

To explore the extent and patterns of agreement, we created the Bland–Altman plot (Fig 1), with 95% statistical limits of agreement (LoA), represented by ± 1.96 standard deviations (SD) of the mean. The mean difference or the bias observed between the methods was 0.04 with a standard deviation of 0.06 and the lower and upper limits of agreement spanning from -0.07 to 0.17. The Bland-Altman Plot can be appreciated visually for any trends of occurrence of data points either equally, above or below the line of mean difference within the limits of agreements. Deming Regression analysis of the values obtained from glass and clot activator vacutainer showed a good correlation (r=0.9) (Fig 2)

**Fig 1:**
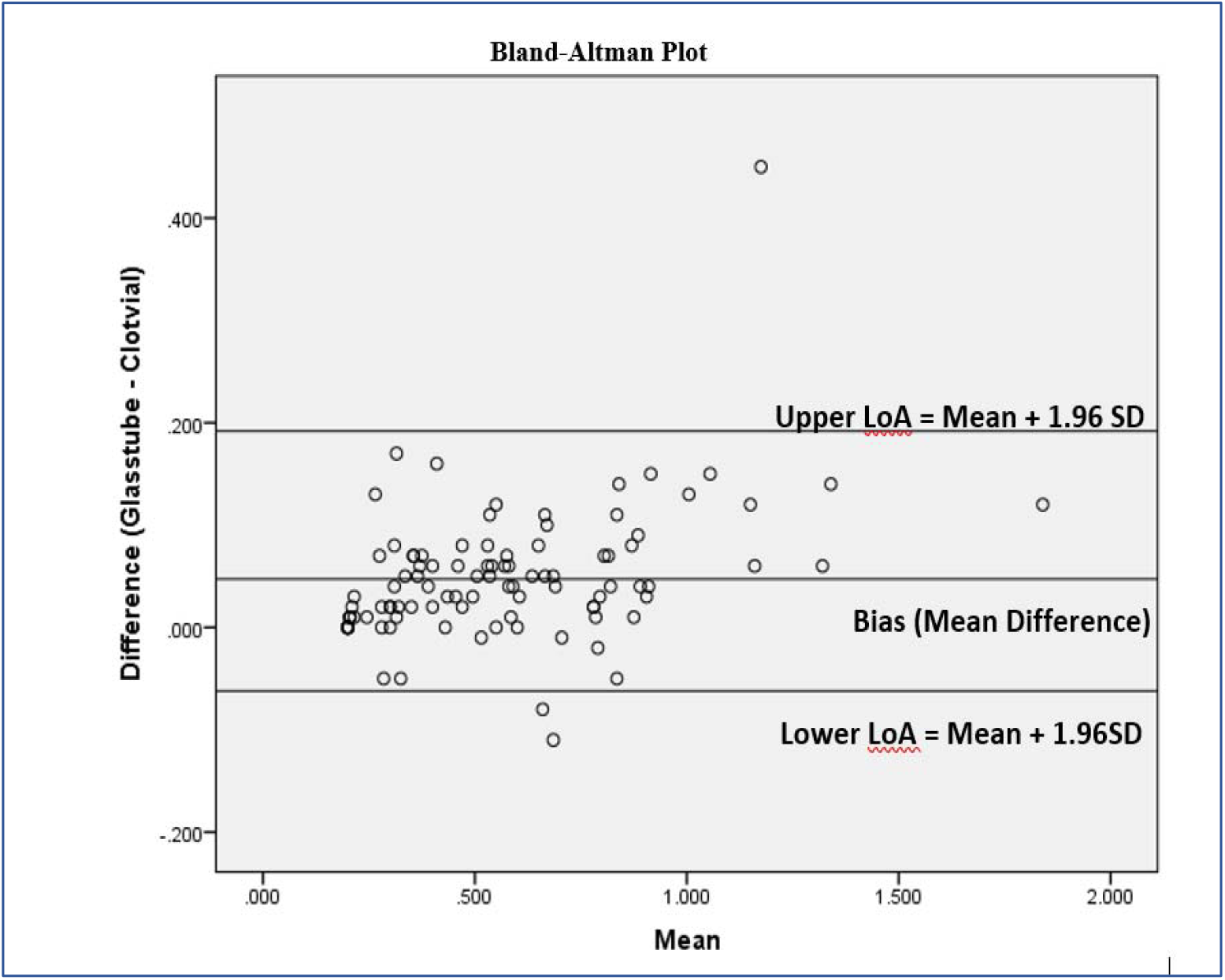
Shows the Bland-Altman Plot of 95% statistical limits of agreement (LoA), represented by ± 1.96 standard deviations (SD) of the mean. The mean difference or the bias observed between the methods was 0.04 with a standard deviation of 0.0648 and the lower and upper limit of agreement spanning from -0.07 to 0.17(mean ±1.96*SD).

**Fig 2:**
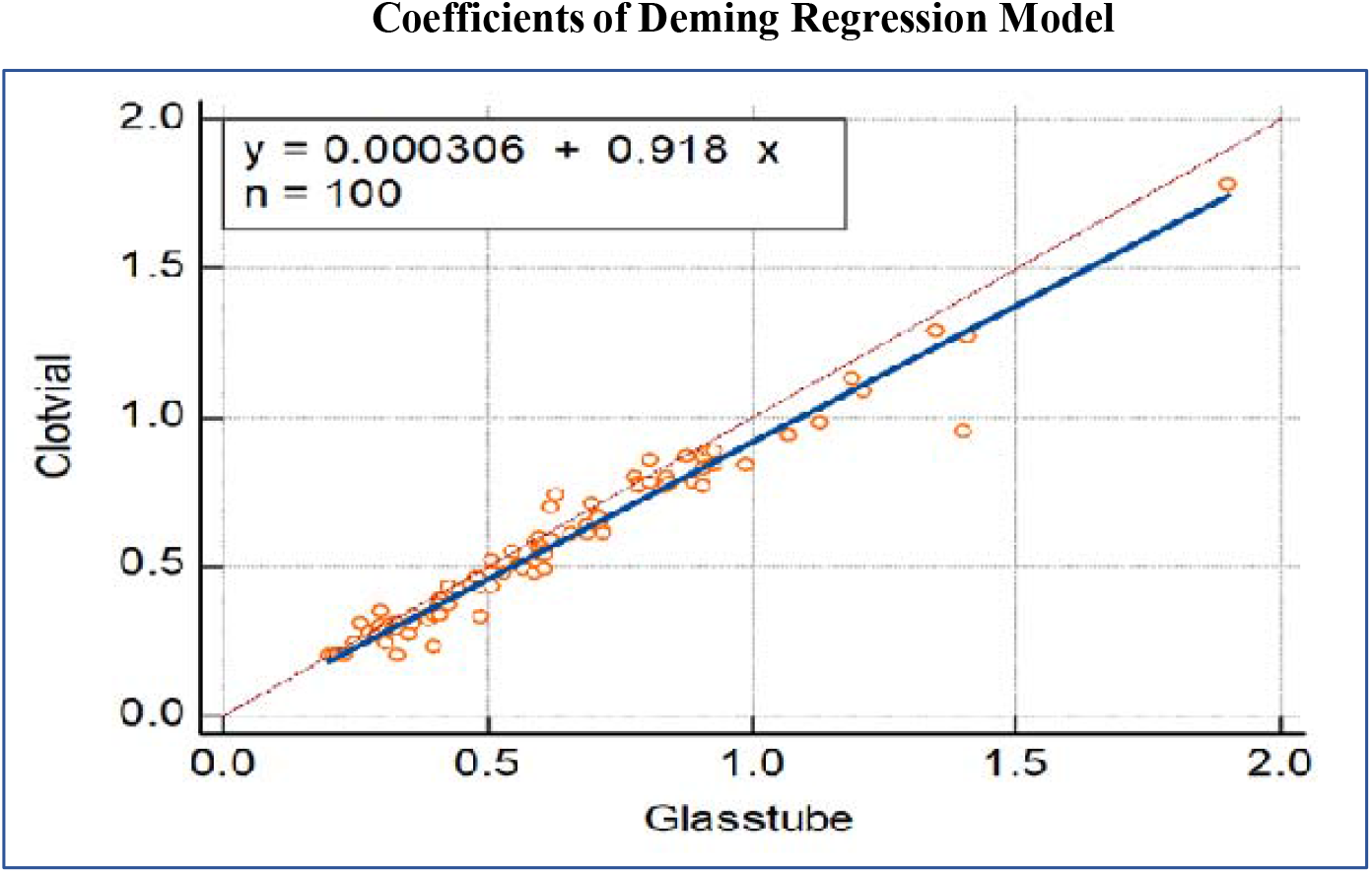
Deming regression showing the error measured during the analysis of values obtained from clot activator vacutainer and glass vial collection tube

To check the stability of the lithium samples in the collection vacutainers (glass vial and clot vacutainer) the samples were further evaluated for lithium levels at 24 hours and 48 hours of the collection which was in addition to the baseline level at 1^st^ hour of collection. The three groups of samples at 1^st^ hour, 24 hours and 48 hours of the collection were tested statistically by one way ANOVA for differences across these groups. For both the Glass vial and Clot vacutainer collection methods (Table 3), there was no statistical difference in the sample groups in different times after collection in ANOVA. The post hoc tukey test was also non-significant for all the groups. Therefore, it can be said that all the groups were not different or same i.e., the lithium levels do not change significantly at 1^st^ hour, 24 hours and 48 hours after collection.

**Table 3:**
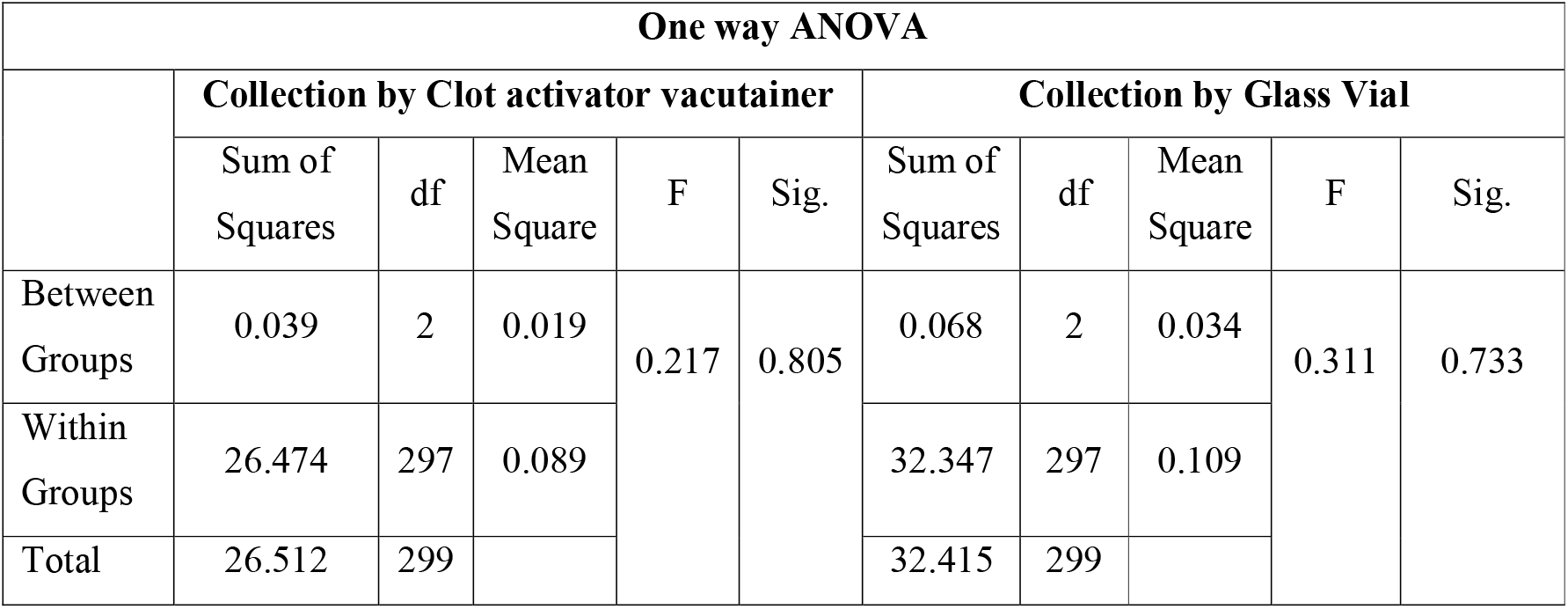
Shows one way ANOVA Analysis done for the lithium values obtained from both the Clot Activator Vacutainer & Glass Vial.

## Discussion

The present era of automation in clinical diagnostics is conveying various analysis (with most of the test parameters of various methodologies) of different individual analysers into a single composite setup. With advanced technologies, the analytical phase of an individual test aims forward to be more accurate and precise. But the preanalytical phase always plays a pivotal role as it drives the analytical phase in the concise path of stringent Quality control Management of the clinical laboratory. The therapeutic drug monitoring for serum lithium is notably challenging for lab experts due to its multi-component dependencies like the patient dosage prescription, last drug intake, time of sample collection and types of collecting vacutainers used in the process of blood sample collection.

Serum lithium, because of its narrow therapeutic range i.e., 0.6-1.2 mmol/L concentration is monitored to ensure patient’s compliance and to avoid lithium overdose thus it is a prerequisite for an individual’s dose adjustment in the treatment of Bipolar mood disorder [7].

In Vitros 4600 autoanalyzer is comprehensive tests facilities where the serum lithium is estimated on dry slide platform assay using the reflectance photometry principle. The VITROS Li slide is a multilayered, analytical element coated on a polyester support. The lithium in the sample is specifically bound by the crown-ether azo reagent (6-dodecyl-6-(2’-hydroxy-5’-(2’’-4’’-dinitrophenylazo) benzyl)-13,13-dimethyl-1,4,8,11-tetraoxacyclotetradecane) and forms a dye complex. As the lithium ion binds to the crown-ether, a shift in the peak absorbance of the dye occurs. The increase in absorbance is proportional to the concentration of lithium in the sample. The intensity of the dye is measured by reflectance spectrophotometry at 600 nm [8,9 10]. The interaction of crown ether with a cation is based on the interaction of charge (the metal cation) and dipole (donor atom within the crown ether ring) which can be influenced by the orientation of the donor atom, cavity size of the crown ether, type of donor atom, the substitution of the electron donating and withdrawing compounds and solvents. [11]

In comparison to common laboratories, serum lithium is estimated mainly by ion selective electrode methods which work mainly on the principle of generation potential difference between the electrodes.[12]

Ikkurthi S *et al* in their study have concluded that plain vacutainers with clot activators show higher bias and statistically significant higher levels of Li than glass vacutainers[13]. Thus, the study further validates the previous study by Sampson *et al*.,1997.[6] that it was shown that silicone present as an additives in the clot activators vacutainer interact with the membrane of Lithium Ion Selective electrodes (ISE) membrane. This interaction results in a false elevation of lithium values when estimated on the ISE analysers.

In our study, both the glass vials and clot activator vacutainers were processed as per the said protocol and found that there is a difference in the value of lithium results. From the literature, Serdarević N *et al* have compare the lithium values obtained from the ISE method with Vitros dry chemistry method and atomic spectrophotometry method, where they have mentioned that there was a positive interference in the ISE method. The Vitros dry slide technology method provides reliable lithium concentration results and is a valid alternative to Atomic Absorption Spectrometry method[14].

We observed that the value of lithium values in the clot activator vacutainer is lower than that of the glass vial. Hence we proved that statistically (refer Table:2). The decreased value of Lithium in the clot activator vacutainers may be due to the presence of silicone additives in the vacutainers which on exposure of the lithium by dry chemistry analyser results in decreased Lithium values. Crown ethers are well known for binding metal cations due to their cyclic cavities and electron donor features. Crown ethers are macrocyclic ligands containing ethyleneoxy units, so called polyethers. They have the ability to crown metal cations. Silicone particles(cations) present in the clot activator vacutainers may have interfered with the crown ethers resulting in decreased value of serum lithium [11].

Vacutainers supplied by different manufacturers vary in the materials and additives that can potentially affect the test performance. Silicone surfactant-coated vacutainers have been shown to interfere with ion-specific electrode measurement of not only lithium but also of ionized magnesium through interaction with ion specific electrode membranes leading to increase in the measured voltage [15].

This confirms that the value obtained from the glass vacutainers is more reliable for patient therapeutic drug monitoring as no such additives are present in the glass vacutainers. In our study, we have also found that there was no statistical difference in the stability of serum lithium values processed from the glass vial and clot activator vacutainer in 1^st^ hour, 24 hours and 48 hours of collection. This might be because of proper storage and processing of the samples at the optimum temperature and the better estimation method as compared to the Ion selective electrode. But Another study has advocated Lithium estimation to be performed within 6 hours as its level falls after that and they have also attributed it to a change in pH affecting the cationic assays [16], which might be one of the causes for the reduced lithium value estimation on different time interval.

## Conclusion

In this study, Lithium levels measured in the clot activated vacutainers were lower as compared to lithium levels measured in the glass vials. There is no apparent change in lithium levels at 1^st^ hour, 24 hours & 48 hours after the collection of blood samples collected in both the glass vacutainer and clot-activated method when measured by the Vitros 4600 dry chemistry analyzer. Collections of blood specimen for lithium estimation should be preferred in glass vial in comparison to the clot activator vacutainer as there was apparently lower Lithium levels due to the interference of silicon particles which is present as an additive in the clot activator vacutainer by its manufacturer for enhancing clot formation process for earlier extraction of serum, within 10-15 minutes of collection of blood.

## Data Availability

All data produced in the present study are available upon reasonable request to the authors

## Author Contribution

All the authors have equally contributed to the accomplishing of the study

